# COVID-19 most vulnerable Mexican cities lack the public health infrastructure to face the pandemic: a new temporally-explicit model

**DOI:** 10.1101/2020.04.10.20061192

**Authors:** Wesley Dáttilo, Alcides Castro e Silva, Roger Guevara, Ian MacGregor Fors, Sérvio Pontes Ribeiro

## Abstract

Recently, a wide array of epidemiological models have been developed to guide public health actors in containing the rapid dissemination of the new severe acute respiratory syndrome coronavirus 2 (SARS-CoV-2), cause of COVID-19. Despite their usefulness, many epidemiological models recently developed to understand the spread of SARS-CoV-2 and infection rates of COVID-19 fall short as they ignore human mobility, limiting our understanding of the spread of the disease, together with the vulnerability of population centers in a broad scale. We developed a new temporally-explicit model and simulated several social distancing scenarios to predict the vulnerability to COVID-19 of 50 Mexican cities that are interconnected by their air transportation network. Additionally, we assessed the sufficiency of the public health infrastructure in the focal cities to face the pandemic over time. Based on our model, we show that the most important cities within the Mexican air transportation network are the most vulnerable to COVID-19, with all assessed public health infrastructure being insufficient to face the modeled scenario for the pandemic after 100 days. Despite these alarming findings, our results show that social distancing could dramatically decrease the total number of infected people (77% drop-off for the 45% distancing scenario when contrasted with no distancing), flattening the growth of infection rate. Thus, we consider that this study provides useful information that may help decision-makers to timely implement health policies to anticipate and lessen the impact of the current pandemic in Mexico.

**Significance Statement:** We used a new temporally-explicit model focused on air transportation networks to predict the vulnerability of 50 focal Mexican cities to COVID-19. We found that most vulnerable cities lack of the required public health infrastructure (i.e., number of inpatient and intensive care unit beds) to face this new pandemic, overloading in all cases after 100 days. However, our results show that a 45% social distancing scenario can reduce the number of infected people by up to 78.7%, flattening the growth rate of people with COVID-19 before infection rates soar exponentially countrywide.

## Introduction

Through history, a number of severe pandemics have shaken humanity without warning (e.g., bulbous plague, smallpox, cholera, Spanish flu). Pandemics are characterized by the spread of an infectious disease that affects a large human population that are not immune to it and have caused millions of human deaths in the past 1,500 years (1, 2). Currently, the world is facing a new pandemic generated by the new severe acute respiratory syndrome coronavirus 2 (SARS-CoV-2), the pathogen behind COVID-19. The virus and its related disease were unknown before the December 2019 outbreak of Wuhan (China). By January 30 2020, the World Health Organization (WHO) declared the COVID-19 situation as a public health emergency of international concern, and 41 days later, in March 11 2020, declared it as a global level pandemic (3). By early April, there were 1.5 million worldwide contagion cases and more than 90,000 deaths (4).

One of the main factors behind the quick dispersion rate of SARS-CoV-2 is its contagion profile. This virus is transmitted through respiratory droplets of infected persons, produced by breathing, sneezing and coughing, and into another individual via mucous membranes (e.g., nose, mouth, and eyes). Thus, the proximity between individuals and their long-distance mobility are two of the main drivers that have been identified in explaining the rapid dispersion of this new airborne disease. Hence, social distancing together with the isolation of infected patients have been suggested as two of the best procedures to decrease infection rates in the absence of a vaccine or other effective treatments (5). In fact, governments from across the globe have promoted lockdowns in the past few weeks. Nonetheless, immerse in globalized practices that demand high mobility of large numbers of people, the daily migration rate across cities, countries, and even continents remains despite the sanitary emergency, adding to the spread of the disease at different spatial scales (6).

Different epidemiological models have been developed for COVID-19 and adapted by to local conditions in order to guide public health actors to contain the rapid advance of this pandemic (7, 8). Most of the existing models, however, do not consider mobility between locations, limiting our understanding on its role in the spatial spread of this highly contagious disease (but see 9, 10). Interestingly, initial COVID-19 cases in most countries were imported cases through international flights. But not only international flights are of concern, as many countries with large territories where domestic flights are the main way between distant locations, are highly important in understanding the spread of the disease. Yet, the importance of airport mobility has been largely overseen by most models, although airports could play a keystone role in the spread of SARS-CoV-2. Thus, including them in further models could allow us to better predict the vulnerability of cities to COVID-19.

In this study, we developed a temporal epidemiological model to predict the vulnerability of 50 cities across Mexico to COVID-19 following the arrival of SARS-CoV-2 to the country. Mexico, with almost 2 million km^2^ of continental territory, is one of the 15 largest countries across the globe, with a population of nearly 120 million inhabitants (11). The first confirmed case of COVID-19 for Mexico was reported on February 28^th^— a case imported from Italy. Forty-five days after the first report, official governmental sources reported more than 3000 confirmed infected patients by the end of the first week of April, with an important spread through community transmission (12). Currently, Mexican cities with more infection cases (i.e., Mexico City, Guadalajara, and Monterrey) have some of the largest and most active international airports in the country, being hubs for both international and domestic flights. Consequently, these cities represent a potential source of transmission of the disease to different regions of the country through the Mexican network of domestic flights. Thus, we consider that it is imperative to understand how COVID-19 could disperse through Mexico’s domestic flight network, assuming cities with a large number of cases as a source of infection to other cities.

The framework of our model is based on a SIR (Susceptible-Infected-Recovered) model and is divided in a metapopulation structure, where 50 focal Mexican cities with airports of national and international category are interconnected by their number of flights. Flights from cities with a high number of confirmed cases of COVID-19 are considered as an additional factor for the spread of the disease in the country, as already described for Brazil (10). In addition to modeling how the COVID-19 will spread across the Mexican territory over time, we consider of crucial importance to know if Mexican cities are prepared with the required healthcare infrastructure to successfully deal with this pandemic under the simulated scenarios. For this, we also assessed the relevant public healthcare infrastructure of the Mexican cities that show to be most vulnerable to COVID-19 by our model. To do so, we calculated the number of infected patients that will need hospitalization on a daily basis. Finally, since social distancing guidelines are being shown to lower the rate of COVID-19 dispersion in many territories, we have also created scenarios of social distancing to simulate its effect on the number of people infected in Mexico over time.

## Materials and Methods

### Susceptible-Infected-Recovered Network (SIR-Net) model

We developed a modified version of the classic discrete Susceptible-Infected-Recovered (SIR; 13) model that runs along cities obeying the connection topology among cities. In the original SIR model, the infection of susceptible individuals occurs given a probability *β* of a healthy being (*S*) encountering an infected one (*I*), also estimating the probability *γ*, at which infected individuals would recover (*R*) from the infection. Analytically:

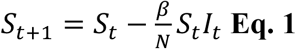

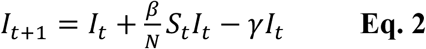

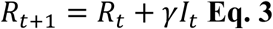

where *t* and *t*+1 represent the current time and a further time, respectively, and *N* = *S* + *I* + *R* is a population constant.

To assess our main goal, we introduced two modifications to the original SIR model in order to simulate the vulnerability of Mexican cities to COVID-19. The first modification refers to the fact that we are applying the model simultaneously among interconnected cities. Thus, our model has *S*^*i*^, *I*^*i*^ and *R*^*i*^, where *i* defines any of the focal cities. We also modified the model to incorporate the spread of the disease under a graph topology defined by regular flights among the 50 focal cities (i.e., representation of a graph in the plane, where the vertices are represented by differing points and the edges) (Code S1). Using the OpenFlights database (14), we were able to track all flights departing and arriving to the airports of the 50 assessed Mexican cities (Dataset S1). Thus, our model (referred to as SIR-Net model hereafter) takes the following form:

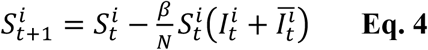

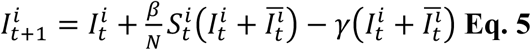

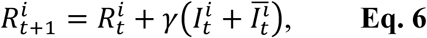

where the upper index *i* indicates the city, and *t* the time (from 0 to 600 days). The term 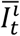 represents the infection added to the *i*^*th*^ city due to the mobility of infected travelers, and it is calculated as follows:

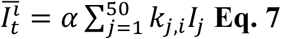

where *k*_*j,i*_ is the number of flights departing from city *i* and arriving to city *j. α* is a newly introduced parameter representing the fraction of traveling infected population. In equation 7 we link equations 4–6 in a network given by *k*_*j,i*_.

To map the spread of COVID-19 in the 50 focal Mexican cities, we initially set a number of infected people in the cities with the highest number of confirmed cases of COVID-19 by early April 2020 (15), as follows: Mexico City: I^22^_0_= 200, Guadalajara: I^15^_0_= 100, and Monterrey: I^26^_0_ = 100. We calibrated our model following a real time series from John Hopkins Coronavirus Research Center (16), which leads to *β* = 0.245 and *γ* = 0.09 (standardized for 242 countries between February 1 and March 31, 2020). The parameter *α* = 0.0001 was estimated using the amount of daily commuting passengers in Mexican airports at the 50 focal cities (17).

### Vulnerability to COVID-19

Based on the results of the SIR-Net model, we assessed the vulnerability of each city to COVID-19. We considered three parameters: (i) the average in the percentage of population affected over the 600 simulated days, (ii) the mean of the daily change in the proportion of the population infected (estimated as: (RI_t+1_ - RI_t_)/(RI_t+1_ + RI_t_), where RI represents the ratio of infected people at a given time t), and (iii) the population size scaled by the natural logarithm, using data from official sources (18). To normalize the final matrix, we divided each cell by the corresponding sum of the column. Afterwards, in the normalized matrix, we the performed a sum of rows and divided them by three. These procedures allowed us to obtain a Vulnerability_index_ for each city over each period of time, where 1 represents maximum vulnerability.

### Estimating the overload of intensive care units

We modeled the time that would take to overload the intensive care units available in the public healthcare system of each focal city under the COVID-19 scenario that assumed no social distancing. Data regarding the existing public healthcare infrastructure was taken from the Mexican Health Secretary (19). This calculation was performed for every simulated day by balancing the number of people that would require an intensive care unit, as well as those that recover or die (estimated by our SIR-Net model). For this case, Δ_N_ represents the variation of the amount of hospitalized patients in a given intensive care unit, in such way that Δ_N_ = (I_1_-I_2_) Δ_t_, where I_1_ is the rate of occupation of intensive care units and I_2_ the rate at which the units are made available either because of recovery or death of patients. Given that the unit of time in our models is the day, Δ_t_ = 1, and Δ_N_ = N_t_+1-N_t_ = I_1_-I_2_, or N_t_+1 = N_t_ + (I_1_-I_2_). In this case, factor I_1_ was calculated as a fraction of infected patients (5%) that would require intensive care units, and to estimate I_2_ and N_t_, we considered an average of 10 days in intensive care for each patient admitted at this level (following 20). Therefore, overloads occur at day t in which N_t_ is higher than the number of intensive care units available in the public healthcare system of a given city.

### Social distancing and the number of infected people

In our SIR-Net model, social distancing was simulated by the decrease of contagion rate (β parameter) given by any method of procedure that directly reduces the potential of an individual to infect another one. Specifically, we tested three hypothetical social distancing scenarios, with contagion decreases of 15%, 30%, and 45%, together with a control with no social distancing. Based on our SIR-Net model, the initial β-value of our model assuming no social distancing was of 0.245. Thus, in our framework, the percentage of social distance directly and proportionally affected further infection rates (Datasets S2-S5). Additionally, since we had already calculated the number of infected people per city 600 days after the initial condition used (Mexico City: I^22^_0_ = 200, Guadalajara: I^15^_0_ = 100, and Monterrey: I^26^_0_ = 100), we estimated the maximum number of people that should be infected in all of the assessed focal cities.

### Data analysis

Initially, we carried out a Spearman rank correlation to assess whether those cities with higher airport closeness centrality (i.e., important cities for connecting different cities within the Mexican air transportation network) were more vulnerable to COVID-19. We calculated closeness centrality (i.e., the reciprocal of the sum of the length of the shortest paths between the node and all other nodes in the network) for each airport using the same air transport network used in our SIR-Net model. We also performed a Spearman correlation to evaluate whether Mexican cities most vulnerable to COVID-19 were related to better public health infrastructure in terms of higher number of inpatient and intensive care beds (including both adult and pediatric beds). The number of beds for each city has was standardized by every 10,000 inhabitants according to the recommendations of the WHO (21). It is notable that our calculations only include Mexican public hospitals (19).

## Results

### Vulnerability of Mexican cities to COVID-19

Using our Net-SIR model, we show that most Mexican cities have a high vulnerability to COVID-19 (Vulnerability_*index*_: 0.86 ± SD 0.02) (Figure 1A). We also found such vulnerability to be positively correlated with airport closeness centrality (rho = 0.86; p <0.001; Figure 1B), indicating that the most important cities in terms of air transportation were the most vulnerable to COVID-19 in our model. Notably, Mexico City and Tijuana showed highest vulnerability values (> 0.9; Dataset S6).

**Figure 1.**
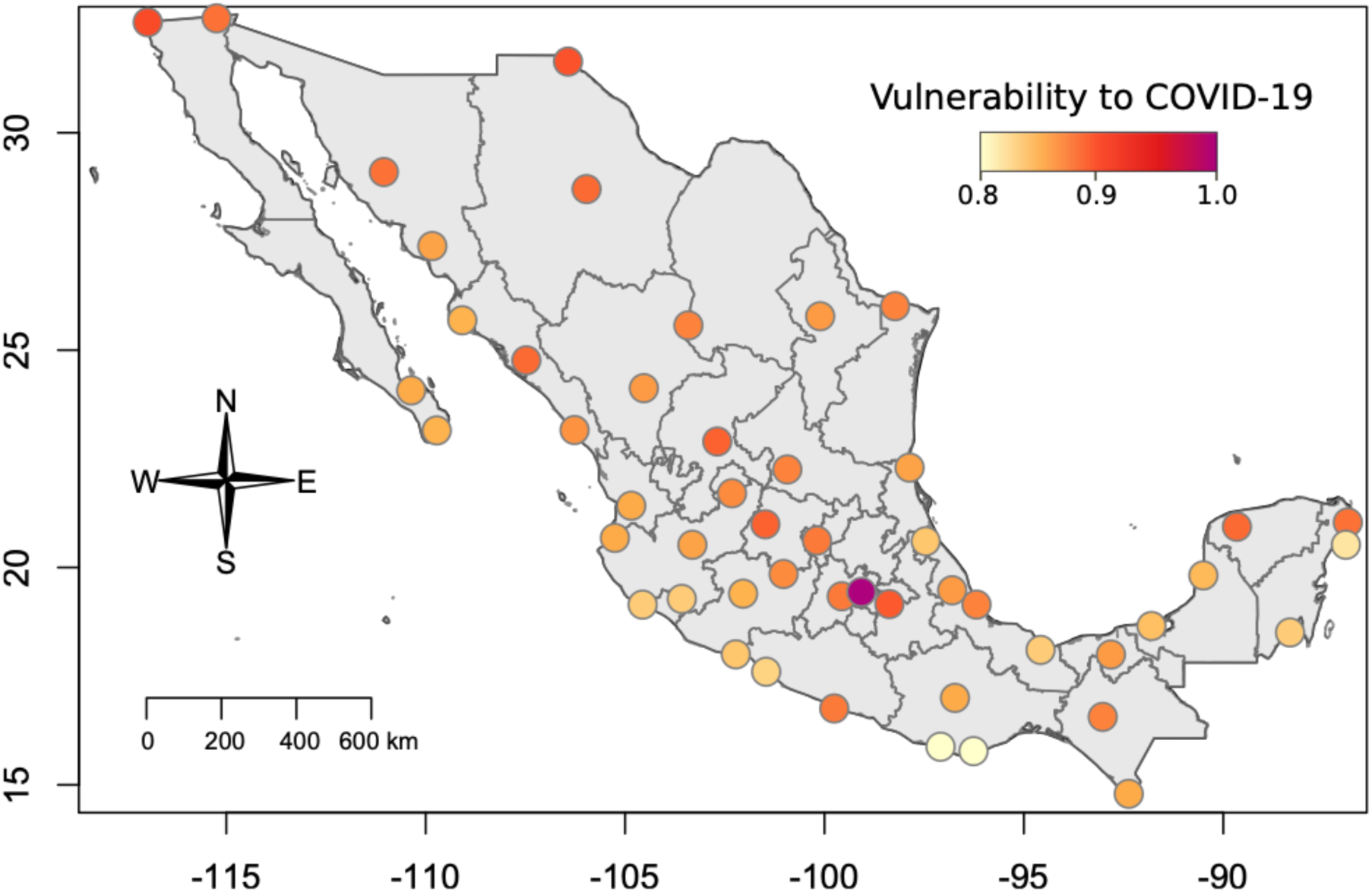
Vulnerability to COVID-19 of 50 Mexican cities with national and international airports and (B) the relationship of its vulnerability with its airport closeness centrality within the Mexican air transportation network.

Regarding our assessment of the potential overload of intensive care units across the focal 50 cities, we found that most of them lack the public health infrastructure to face the pandemic. Our results are clear in showing that cities that were identified to be more vulnerable to COVID-19 do not have a greater number of inpatient (rho = −0.21; p = 0.16. Figure 2A) nor intensive care units (rho = 0.26; p = 0.06; Figure 2B) for every 10,000 inhabitants. Most importantly, we found that all cities would overload their intensive care system before 100 days after the initial condition of our model (66.4 ± SD 25.7 days; Figure 3).

**Figure 2.**
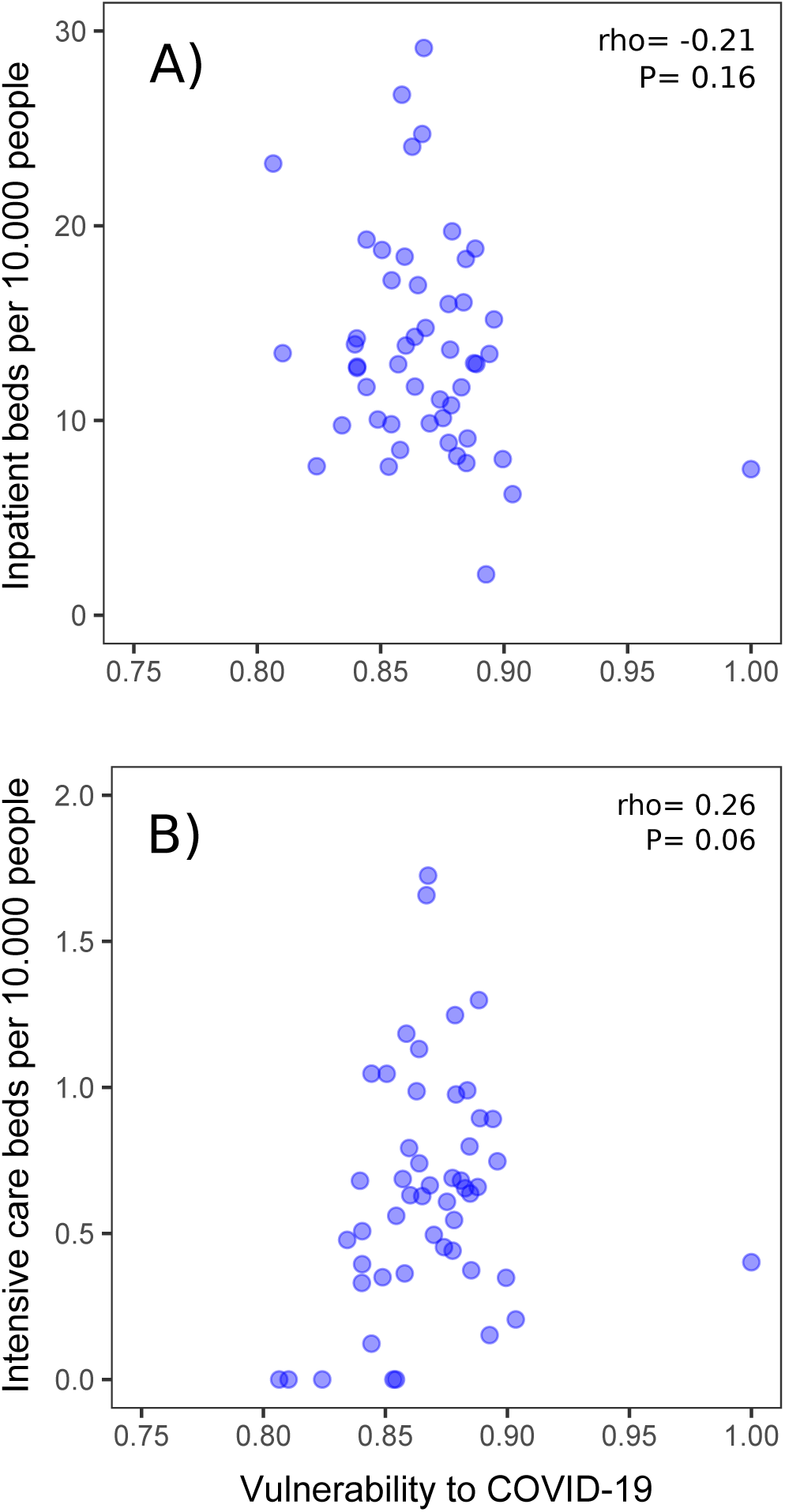
Relationship between the vulnerability of cities to COVID-19 and A) inpatient and B) intensive care beds per 10,000 people.

**Figure 3.**
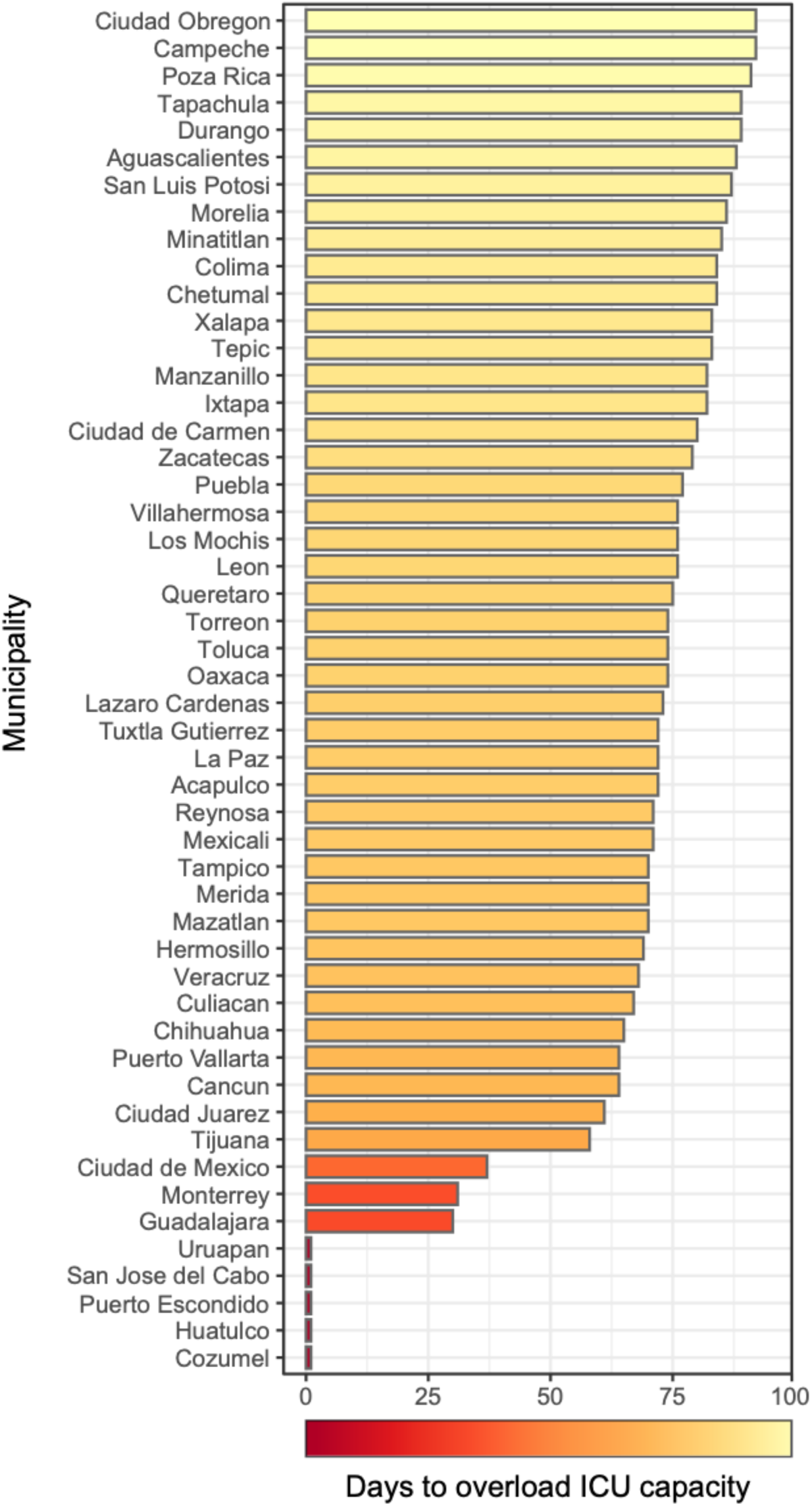
Days to overload the intensive care units (ICU) in the 50 Mexican cities studied according to our SIR-Net model.

**Figure 4.**
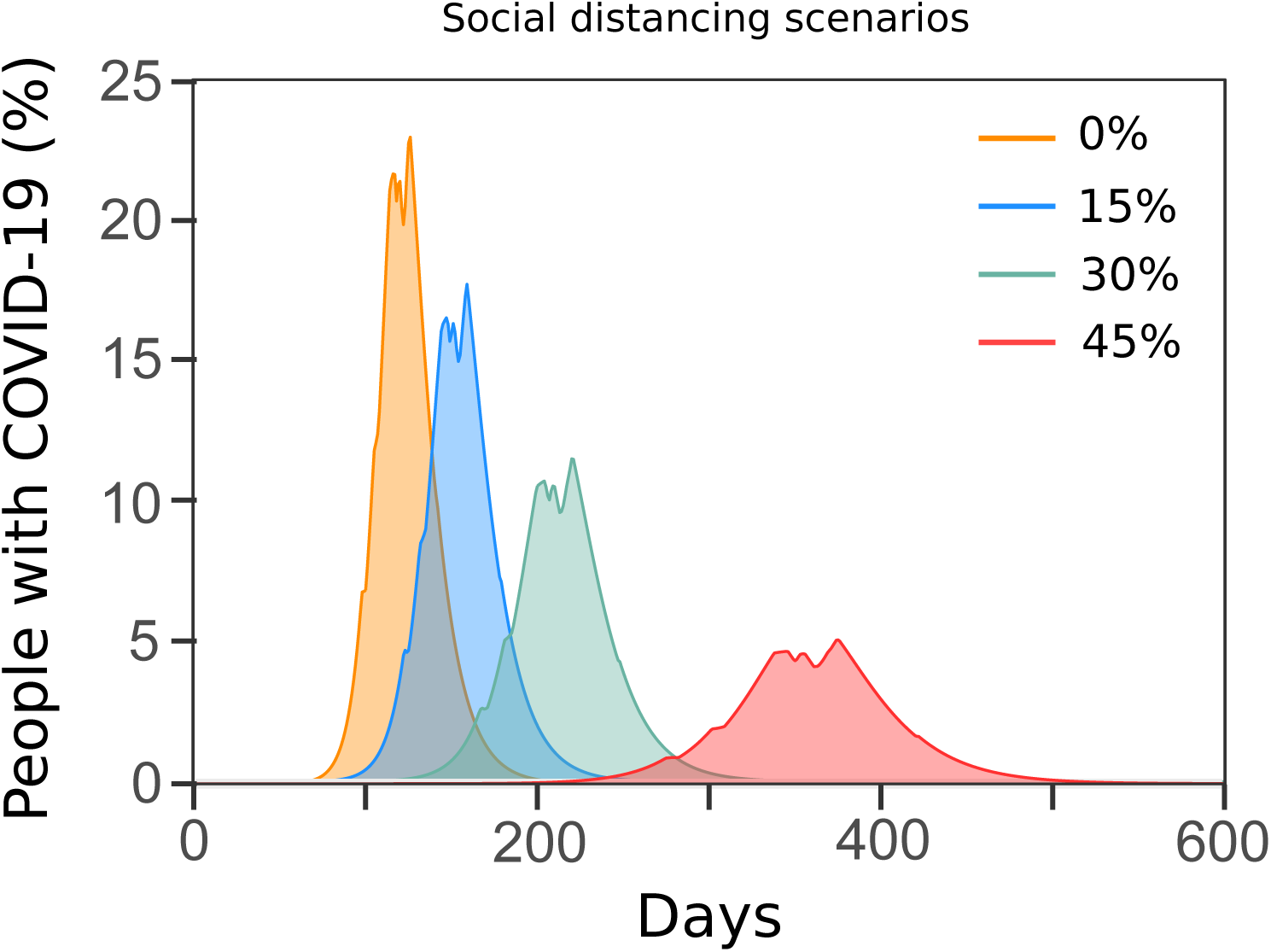
Median percentage of people infected with COVID-19 considering the 50 cities studied over 600 days with different social distancing scenarios (0%, 15%, 30% and 45%). Estimation based on our Network Susceptible-Infected-Recovered (SIR-Net) model.

### Scenarios of social distancing

When we modeled the behavior of the infection rate and spread of COVID-19 over time without social distancing, we observed that the percentage of infected people in the 50 focal cities sky-rocketed swiftly between days 83 to 172, reaching its peak close to day 126, with 22.9% of the population being infected (13,099,464 infected people). When we applied measures of social distancing in our SIR-Net model, we found significant and progressive changes in the number of people infected over time. In the 15% social distancing scenario, we observed that the percentage of infected people rose, again quickly, between days 107 and 209, to a maximum peak in day 159, with an overall 17.7% of the population of the 50 focal cities be infected (10,103,144 infected people). For the 30% of social distancing scenario, we observed that the percentage of infected people in the 50 focal cities rose, less steeply, between days 153 and 276, to a maximum peak in day 220, with 11.4% of the population infected (6,692,639 infected people). Yet, we detected an impressive decrease in the rate of infection (i.e., flattening of the curve) in the 45% social distancing scenario. The percentage of infected people in the country in that last scenario would start to rise between days 284 and 436, to a maximum peak around day 375, with only 5.1% of the assessed population infected (3,039,553 infected people).

## Discussion

Using a tailored modification of a SIR model based on the transportation of people between 50 focal Mexican cities, we show that the most important Mexican cities in terms of their air transportation network (with higher closeness centrality) are the most vulnerable to COVID-19. Woefully, most vulnerable cities lack of the required public health infrastructure (i.e., number of inpatient and intensive care unit beds) to face this pandemic considering the simulated infection numbers. Interestingly, when we simulated the effect of social distancing with our model, it shows that 45% of social distancing could decrease up to 78.7% the number of infected people, flattening the growth rate of people with COVID-19 before infection rates soar exponentially.

Using the proposed SIR-Net model, we show that the most important cities within the Mexican air transportation network are the most vulnerable to COVID-19. Recently, Ribeiro et al (10) showed similar results for the modeled expansion of SARS-CoV-2 in Brazil, with airport closeness centrality related to COVID-19 model severeness. Previous studies have shown that cities that serve as transfer points (i.e., airport hubs) are important drivers for the spread of diseases across the world. Indeed, the John Hopkins Coronavirus Research Center and other several monitors of the spread of SARS-CoV-2 across the world have shown that the critical starting points of the pandemic in many countries are airport hubs, such as Milan, Rome, Madrid, and New York. Results of this study together with previous knowledge suggest that controlling air traffic in key cities, mostly those with high airport closeness centrality, could mold globalized pandemics such as COVID-19. Thus, the implementation of timely airport sanitarian control could have slowed down the spreading of COVID-19 in countries with a massive flow of flight routes—a big lesson for future control of infections outbreaks.

Regarding Mexican city vulnerability, our results clearly indicate that Monterrey, Guadalajara, and Mexico City are the most vulnerable cities to COVID-19. These Mexican cities are the most economically important urban centers in the country, are the most populated, and are directly connected with many cities from across the world through their international airports. According to our predictions, if no social distancing is assured, these cities together could add up to 13 million infected inhabitants with COVID-19 over the course of a year (Dataset S6). Thus, based on our model, we suggest that considerable efforts should be focused in these cities in order to control and monitor the spread of COVID-19. These cities have great potential to a large number of infected people and also could serve as a source of spread of the disease to other cities and regions of the country, and even playing a crucial role in further outbreaks of COVID-19. In relation to the capacity of the current infrastructure required to face this pandemic in the 50 assessed focal cities, our results show that once COVID-19 patients demand intensive care units, it would take less than 100 days to overloaded the capacity of their public healthcare systems. Additionally, the most vulnerable cities to COVID-19 identified by our model do not have a higher number of inpatient and intensive care unit beds when standardized for every 10,000 inhabitants. If the number of infected people continues to grow in the assessed Mexican cities as it did in Wuhan (China), for instance, no Mexican city would be prepared to deal with this pandemic. The latter is due to the estimate of 24.5 inpatient beds and 2.6 intensive care unit beds for every 10,000 people would be needed during the peak of the epidemic (21). Among the focal assessed Mexican cities, only Villahermosa, La Paz, and Monterrey have a sufficient number of inpatient beds than the estimated ones that would be needed per 10,000 (i.e., 29.1, 26.7 and 24.7, respectively). However, no Mexican city has enough intensive care unit beds, all being below 1.8 intensive care unit beds per 10,000 people, reason why they would overload 100 days after the initial condition of our model.

One of the control measures that appear to be slowing the spread of coronavirus in some regions is the social distancing, albeit its well-discussed social and economic ramifications (22). We here show that 45% social distancing scenario decreases the modeled number of infected people from 13,099,464 to 3,039,553, representing not only a 78.7% drop-off, but also a flattening of the contagion growth rate, and thus not overburdening the public healthcare system. These results support the suggestion of the Mexican Sub-secretary of Prevention and Health Promotion for people to stay home and only going out for food and medicine supply, and in case of emergencies, always following social distancing and hygiene measures suggested by the WHO.

The model proposed in this paper considers some key aspects of the dissemination of COVID-19 among 50 focal Mexican cities, hoping that it adds to the existing knowledge and reverberates into public actions. Thus, we consider that it is important to highlight some of the main limitations of our model. First, we only consider air transport to estimate the mobility of people between cities; however, the we are well aware that the Mexican road network is one important means of communication, representing an additional potential driver for the spatial spread of the disease. Second, our model only considers the health infrastructure in terms of inpatient and intensive care unit beds from the public healthcare network. Most recently, the Mexican Government increased hospital capacity in some cities, which could mold the outcome of our results if considered in the model. Third, there are additional factors that have been identified as important in facing the spread of COVID-19 (e.g., protective equipment, medical personnel, and mechanical ventilators; 23) that are not considered in our model. Finally, we assessed social distancing as a general grouping factor behind the decrease of contagion rates, which could not only include well known specific drivers of such phenomenon (e.g., thorough hand washing, staying at home, keeping distance when leaving home). Taking into account all of the limitations of our model and the available data used to construct it, we show how the Mexican air transport network could increase the vulnerability of cities to COVID-19. Also, our simulations suggest that the available public healthcare infrastructure in the most vulnerable studied focal cities is very likely to be insufficient to face this pandemic. Despite most scenarios derived from our model are not optimistic for the coming months, our model clearly shows that social distancing is a remarkable measure to control the growth rate of total COVID-19 cases before infection rates soar exponentially in the country. We observed important differences between 0–30% social distancing scenarios and that of 45%, where the curve is clearly flattened. Altogether, our findings for 50 evaluated focal cities add to our knowledge in understanding the potential future behavior of COVID-19 in Mexico. We hope that our model and results provide useful information for governmental agencies, at their different levels, in the mitigation of the impacts of this pandemic.

## Data Availability

All data, including the model code is available in the supplementary material

## Acknowledgments

We also thank Gabriel Alfonso Carranco Sapiéns for his help in finding information on Mexican hospital infrastructure. The authors also thank Jorge Soberón for encouraging the writing of this manuscript. SPR is granted by CNPq-Brazil.

## Figure captions

**Figure 4.** Median percentage of people infected with COVID-19 considering the 50 cities studied over 600 days with different social distancing (SD) scenarios (0%, 15%, 30% and 45%). Estimation based on our SIR-Net model.

